# Neutralization of SARS-CoV-2 Omicron pseudovirus by BNT162b2 vaccine-elicited human sera

**DOI:** 10.1101/2021.12.22.21268103

**Authors:** Alexander Muik, Bonny Gaby Lui, Ann-Kathrin Wallisch, Maren Bacher, Julia Mühl, Jonas Reinholz, Orkun Ozhelvaci, Nina Beckmann, Ramón de la Caridad Güimil Garcia, Asaf Poran, Svetlana Shpyro, Hui Cai, Qi Yang, Kena A. Swanson, Özlem Türeci, Ugur Sahin

**Affiliations:** BioNTech, An der Goldgrube 12, 55131 Mainz, Germany; BioNTech US; 40 Erie Street, Cambridge, MA, 02139, USA; Pfizer, 401 N Middletown Rd., Pearl River, NY 10960, U.S.A.; HI-TRON – Helmholtz Institute for Translational Oncology Mainz by DKFZ, Obere Zahlbacherstr. 63, 55131 Mainz, Germany; TRON gGmbH – Translational Oncology at the University Medical Center of the Johannes Gutenberg, University Freiligrathstraße 12, 55131 Mainz, Germany

## Abstract

A new severe acute respiratory syndrome coronavirus 2 (SARS-CoV-2) lineage, B.1.1.529, was recently detected in Botswana and South Africa and is now circulating globally. Just two days after it was first reported to the World Health Organization (WHO), this strain was classified as a variant of concern (VOC) and named Omicron. Omicron has an unusually large number of mutations, including up to 39 amino acid modifications in the spike (S) protein, raising concerns that its recognition by neutralizing antibodies from convalescent and vaccinated individuals may be severely compromised. In this study, we tested pseudoviruses carrying the SARS-CoV-2 spike glycoproteins of either the Wuhan reference strain, the Beta, the Delta or the Omicron variants of concern with sera of 51 participants that received two doses or a third dose (≥6 months after dose 2) of the mRNA-based COVID-19 vaccine BNT162b2. Immune sera from individuals who received two doses of BNT162b2 had more than 22-fold reduced neutralizing titers against the Omicron as compared to the Wuhan pseudovirus. One month after a third dose of BNT162b2, the neutralization titer against Omicron was increased 23-fold compared to two doses and antibody titers were similar to those observed against the Wuhan pseudovirus after two doses of BNT162b2. These data suggest that a third dose of BNT162b2 may protect against Omicron-mediated COVID-19, but further analyses of longer-term antibody persistence and real-world effectiveness data are needed.

## Main Text

Since the first reports of severe acute respiratory syndrome coronavirus 2 (SARS-CoV-2) in humans in December 2019, numerous genetically distinct lineages have evolved. Among those, variants of concern (VOC), especially Alpha and Delta, were associated with increased transmissibility and sparked new waves of infection, with Delta, first designated a VOC on 11^th^ May-2021 (*1*), quickly becoming the globally dominant variant (*2*). On November 26^th^ 2021, a new VOC – Omicron – was reported by the World Health Organization (WHO) (*3*). Omicron is a highly divergent variant and harbors a hitherto unprecedented number of mutations in its spike (S) glycoprotein (*4*). 15 mutations are located in the receptor-binding domain (RBD) and another eight mutated sites are found in the N-terminal domain (NTD), both being immunodominant targets of neutralizing antibodies elicited by COVID-19 vaccines or by SARS-CoV-2 infection (*5, 6*). Some amino acid changes (Δ69/70, T95I, G142D, Δ145, K417N, T478K, N501Y, and P681H) are shared mutations also found in the Alpha, Beta, Gamma or Delta VOCs and were described to lead to increased transmissibility, and to a typically mild partial escape from vaccine-induced humoral immunity (*7–10*).

The BNT162b2 COVID-19 vaccine contains lipid nanoparticle formulated mRNA that encodes the SARS-CoV-2 spike glycoprotein from the parental Wuhan reference strain (*11*). Administration of two 30-μg doses of BNT162b2 was shown to have 95% efficacy in the pivotal Phase III trial (*12*), and shown to elicit strong antibody responses effectively neutralizing the parental strain as well as diverse SARS-CoV-2 variants of concern (*13–15*). The effect of many of the new mutations in Omicron on recognition by neutralizing antibodies in convalescent and vaccinated individuals is unknown, as is their effect upon vaccine effectiveness.

To evaluate whether BNT162b2-elicited antibodies (*11*) are capable of neutralizing the Omicron variant, we used a pseudovirus neutralization test (pVNT), an assay system that has been shown to be in close concordance with live SARS-CoV-2 neutralization assays (*16, 17*). We generated vesicular stomatitis virus (VSV)-SARS-CoV-2-S pseudoviruses bearing the spike proteins of either the Wuhan strain, Omicron, Beta (as a benchmark for partially reduced neutralization (*7*) without major impact on effectiveness (*18, 19*)) or Delta (the currently predominant strain as of December 2021). BNT162b2 immune sera from individuals of 20-72 years of age (with over one third 56 years of age and older, Table S1) were obtained from different clinical studies – the Phase 1/2 trial BNT162-01 (NCT04380701) and the Phase 2 rollover trial BNT162-14 (NCT04949490), conducted in Germany; and the global Phase 2 trial BNT162-17 (NCT05004181) (see Methods). Neutralizing titers against VSV-SARS-CoV-2-S pseudoviruses were analyzed with serum drawn from 32 participants in the BNT162-01 trial 21 days (median of 22 days; range 19-23 days) after two doses of BNT162b2, and with serum drawn from 30 participants in the BNT162-14 (n=11) and BNT162-17 (n=19) trials at 1 month (median of 28 days; range 26-30 days) after the third dose of BNT162b2. Nine individuals included in this analysis were rolled over from the BNT162-01 into the BNT162-14 trial.

After two doses of BNT162b2, geometric mean neutralization titers (GMT) against Omicron pseudovirus were 22.8-fold lower compared to the Wuhan reference pseudovirus (Fig. 1; GMT of 7 vs. 160). 20 out of 32 immune sera displayed no detectable neutralizing activity against Omicron (Table S2). In contrast, the majority of sera neutralized Beta and Delta pseudoviruses with GMTs of 24 and 73, respectively. This corresponds to a 6.7-fold and 2.2-fold reduction in neutralization activity compared to the Wuhan pseudovirus and is in line with previous reports (*11, 14, 15, 20*).

**Fig. 1.**
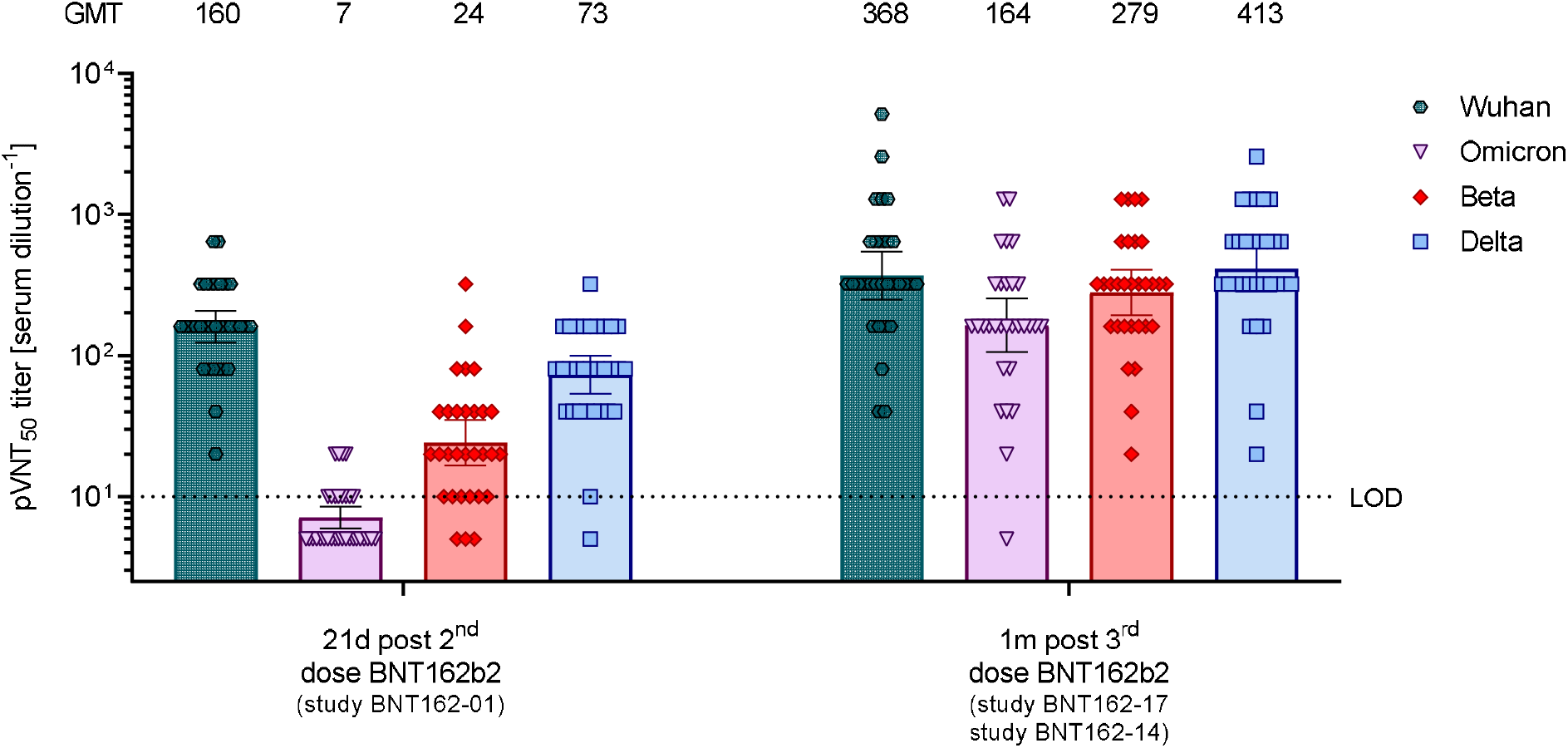
50% pseudovirus neutralization titers (pVNT_50_) of sera from vaccine recipients collected after two or three doses of BNT162b2 against VSV-SARS-CoV-2-S pseudovirus bearing the Wuhan, Omicron, Beta or Delta variant spike protein. N=32 sera from participants in study BNT162-01 drawn at 21 days after dose 2, and n=30 sera from participants in the BNT162-14 (n=11) and BNT162-17 trials (n=19) drawn at 1 month after dose 3 were tested. Each serum was tested in duplicate and geometric mean 50% pseudovirus neutralizing titers (GMTs) were plotted. For values below the limit of detection (LOD), LOD/2 values were plotted. Group GMTs (values) and 95% confidence intervals are indicated.

One month after the third BNT162b2 dose, neutralizing GMTs against the Omicron variant pseudovirus increased 23.4-fold compared to neutralizing GMTs at 21 days after the second dose (GMT of 164 vs. 7); achieving titers comparable to the neutralization against the reference Wuhan pseudovirus at 21 days after two doses of BNT162b2 (GMT of 164 vs. 160). 29 of the 30 sera were capable of neutralizing the Omicron pseudovirus (Table S3). The third dose of BNT162b2 also increased neutralizing activity against Beta, Delta and Wuhan pseudoviruses, with GMTs of 279, 413, and 368, respectively.

BNT162b2 vaccination induces strong poly-epitopic T cell responses, directed against multiple epitopes spanning the length of the spike protein (*11*). To assess the risk of immune evasion of CD8^+^ T cell responses by Omicron, we investigated a set of HLA class I restricted T cell epitopes from the Wuhan spike protein sequence that were reported in the Immune Epitope Database to be immunogenic (IEDB, n=244; see Methods). Despite the multitude of mutations in the Omicron spike protein, 85.3% (n=208) of the described class I epitopes were not impacted on the amino acid sequence level, indicating that the targets of the vast majority of T cell responses elicited by BNT162b2 may still be conserved in the Omicron variant (fig. S1).

In summary, our data indicate that two doses of BNT162b2 may not be sufficient to protect against infection with the Omicron variant. The observed 22.8-fold reduction in neutralizing activity for immune sera drawn 21 days after the primary 2-dose series of BNT162b2 confirms preliminary reports describing a 20- to 40-fold reduction in titers (*21, 22*). A third dose of BNT162b2 boosts Omicron neutralization capability to a level similar to that observed after two doses against the Wuhan pseudovirus. Similar results have been reported for subjects vaccinated with BNT162b2 and previously infected by the ancestral SARS-CoV-2 strain (*21*). Our observation that a third dose augments antibody-based immunity against Omicron (and also against Beta and Delta VOC pseudovirus) is in line with previous observations that a third vaccination broadens humoral immune responses against VOCs (*23*).

The analysis presented here has evaluated and compared serum panels from different studies with a limited sample size, with the timing of the third dose not consistent between participants. Future analyses will evaluate antibody persistence.

Neutralizing antibodies represent a first layer of adaptive immunity against COVID-19. T cell responses play a vital role as a second layer of defense, in particular in the prevention of severe COVID-19 (*24*). CD8^+^ T cell responses in individuals vaccinated with BNT162b2 are polyepitopic (*11*), and our analyses suggest that CD8^+^ T cell recognition of Omicron spike glycoprotein epitopes are largely preserved. Our data show that a third BNT162b2 dose effectively neutralizes Omicron pseudovirus at similar levels observed after two doses of BNT162b2 against wild-type Wuhan pseudovirus. Early reports estimate moderate to high vaccine effectiveness against symptomatic infection due to Omicron; 70 to 75% has been reported from the UK shortly after the booster dose (*25, 26*). Further clinical trial and real world data will soon emerge to address the effectiveness of a third dose with BNT162b2 against COVID-19 mediated by Omicron.

## Data Availability

Materials are available from the authors under a material transfer agreement with BioNTech.

## Acknowledgments

We thank the BioNTech German clinical Phase 1/2 trial (NCT04380701, EudraCT: 2020-001038-36), the German Phase 2 rollover booster trial (NCT04949490, EudraCT: 2021-002387-50) and the global clinical Phase 2 trial (NCT04380701) participants, from whom the post-immunization human sera were obtained. We thank the many colleagues at BioNTech and Pfizer who developed and produced the BNT162b2 vaccine candidate. We thank S. Jägle for logistical support, and A. Finlayson for editorial and medical writing support. We thank Christina Heiser, Ayca Telorman, Kimberly Krüger, Claudia Müller, Amy Wanamaker, Nicki Williams and Jennifer VanCamp for sample demographics support.

## Author contributions

U.S., Ö.T., and A.M. conceived and conceptualized the work. A.M, B.G.L., J.R., H.C., Q.Y., K.A.S and R.G planned and supervised experiments. A.M., A.W., B.G.L, J.M., J.R., M.B., N.B. and R.G. performed experiments. A.M., A.P., B.G.L., J.R., K.A.S., O.O., R.G., and S.S. and analyzed data. U.S., Ö.T., A.M., and K.A.S. interpreted data and wrote the manuscript. All authors supported the review of the manuscript.

## Data and materials availability

Trial participant baseline characteristics are provided in table S1. The neutralization titers are provided in table S2 and table S3. Materials are available from the authors under a material transfer agreement with BioNTech.

## Supplementary Materials

Materials and Methods

Figs. S1

Tables S1-S3

References

## References

1. World Health Organization, Tracking SARS-CoV-2 variants (available at https://www.who.int/en/activities/tracking-SARS-CoV-2-variants/).

2. World Health Organization, SARS-CoV-2 Delta variant now dominant in much of European region; efforts must be reinforced to prevent transmission, warns WHO Regional Office for Europe and ECDC (available at https://www.euro.who.int/en/media-centre/sections/press-releases/2021/sars-cov-2-delta-variant-now-dominant-in-much-of-european-region-efforts-must-be-reinforced-to-prevent-transmission,-warns-who-regional-office-for-europe-and-ecdc).

3. WHO Technical Advisory Group on SARS-CoV-2 Virus Evolution (TAG-VE), Classification of Omicron (B.1.1.529): SARS-CoV-2 Variant of Concern (2021).

4. WHO Headquarters (HQ), WHO Health Emergencies Programme, Enhancing Readinessfor Omicron (B.1.1.529): Technical Brief and Priority Actions for Member States (2021).

5. L. Premkumar et al., The receptor binding domain of the viral spike protein is an immunodominant and highly specific target of antibodies in SARS-CoV-2 patients. Science immunology. 5 (2020), doi:10.1126/sciimmunol.abc8413.

6. W. T. Harvey et al., SARS-CoV-2 variants, spike mutations and immune escape. Nature reviews. Microbiology. 19, 409–424 (2021), doi:10.1038/s41579-021-00573-0.

7. P. Wang et al., Antibody resistance of SARS-CoV-2 variants B.1.351 and B.1.1.7. Nature. 593, 130–135 (2021), doi:10.1056/NEJMc2031364.

8. D. Planas et al., Reduced sensitivity of SARS-CoV-2 variant Delta to antibody neutralization. Nature. 596, 276–280 (2021), doi:10.1038/s41586-021-03777-9.

9. Q. Wang et al., Functional differences among the spike glycoproteins of multiple emerging severe acute respiratory syndrome coronavirus 2 variants of concern. iScience. 24, 103393 (2021), doi:10.1016/j.isci.2021.103393.

10. A. J. Greaney et al., Complete Mapping of Mutations to the SARS-CoV-2 Spike Receptor-Binding Domain that Escape Antibody Recognition. Cell host & microbe. 29, 44 (2021), doi:10.1016/j.chom.2020.11.007.

11. U. Sahin et al., BNT162b2 vaccine induces neutralizing antibodies and poly-specific T cells in humans. Nature. 595, 572–577 (2021), doi:10.1038/s41586-021-03653-6.

12. F. P. Polack et al., Safety and Efficacy of the BNT162b2 mRNA Covid-19 Vaccine. The New England journal of medicine. 383, 2603–2615 (2020), doi:10.1056/NEJMoa2034577.

13. A. Muik et al., Neutralization of SARS-CoV-2 lineage B.1.1.7 pseudovirus by BNT162b2 vaccine-elicited human sera. Science (New York, N.Y.). 371, 1152–1153 (2021), doi:10.1126/science.abg6105.

14. J. Liu et al., BNT162b2-elicited neutralization of B.1.617 and other SARS-CoV-2 variants. Nature. 596, 273–275 (2021), doi:10.1038/s41586-021-03693-y.

15. Y. Liu et al., Neutralizing Activity of BNT162b2-Elicited Serum. The New England journal of medicine. 384, 1466–1468 (2021), doi:10.1056/NEJMc2102017.

16. J. B. Case et al., Neutralizing Antibody and Soluble ACE2 Inhibition of a Replication-Competent VSV-SARS-CoV-2 and a Clinical Isolate of SARS-CoV-2. Cell host & microbe. 28, 475–485.e5 (2020), doi:10.1016/j.chom.2020.06.021.

17. A. B. Vogel et al., BNT162b vaccines protect rhesus macaques from SARS-CoV-2. Nature. 592, 283–289 (2021), doi:10.1038/s41586-021-03275-y.

18. L. J. Abu-Raddad, H. Chemaitelly, A. A. Butt, Effectiveness of the BNT162b2 Covid-19 Vaccine against the B.1.1.7 and B.1.351 Variants. The New England journal of medicine. 385, 187–189 (2021), doi:10.1056/NEJMc2104974.

19. O. Mor et al., BNT162b2 vaccine effectiveness was marginally affected by the SARS-CoV-2 beta variant in fully vaccinated individuals. Journal of clinical epidemiology. 142, 38–44 (2021), doi:10.1016/j.jclinepi.2021.10.011.

20. A. Kuzmina et al., SARS CoV-2 Delta variant exhibits enhanced infectivity and a minor decrease in neutralization sensitivity to convalescent or post-vaccination sera. iScience. 24, 103467 (2021), doi:10.1016/j.isci.2021.103467.

21. S. Cele et al., SARS-CoV-2 Omicron has extensive but incomplete escape of Pfizer BNT162b2 elicited neutralization and requires ACE2 for infection. medRxiv : the preprint server for health sciences (2021), doi:10.1101/2021.12.08.21267417;

22. A. Wilhelm et al., Reduced Neutralization of SARS-CoV-2 Omicron Variant by Vaccine Sera and Monoclonal Antibodies (2021).

23. A. R. Falsey et al., SARS-CoV-2 Neutralization with BNT162b2 Vaccine Dose 3. The New England journal of medicine. 385, 1627–1629 (2021), doi:10.1056/NEJMc2113468.

24. A. Bertoletti, N. Le Bert, M. Qui, A. T. Tan, SARS-CoV-2-specific T cells in infection and vaccination. Cellular & molecular immunology. 18, 2307–2312 (2021), doi:10.1038/s41423-021-00743-3.

25. UK Health Security Agency, SARS-CoV-2 variants of concern and variants under investigation in England - Technical briefing 31 (available at https://assets.publishing.service.gov.uk/government/uploads/system/uploads/attachment_data/file/1040076/Technical_Briefing_31.pdf).

26. N. Ferguson et al., Report 49: Growth, population distribution and immune escape of Omicron in England (2021) (available at https://www.imperial.ac.uk/media/imperial-college/medicine/mrc-gida/2021-12-16-COVID19-Report-49.pdf).

