## Supplementary for "Neutralization of SARS-CoV-2 Omicron pseudovirus by BNT162b2 vaccine-elicited human sera"

BNT162b2 vaccine-elicited human sera

**This PDF file includes:**

Materials and Methods

Fig. S1

Tables S1 to S3

**Materials and Methods**

Clinical Studies

This research used samples from 18-85 year old participants of the German Phase 1/2 trial BNT162-01 (NCT04380701) vaccinated with 2-dose primary series 30 µg BNT162b2 with a 21 ± 2 d dosing interval, that has been described elsewhere (*1*). In addition, samples were used from participants in subgroups of two ongoing clinical trials, BNT162-14 (NCT04949490) aged 18-85 and BNT162-17 (NCT05004181) aged 18-55 years old.

Participants of BNT162-01 who received the 2-dose primary series with BNT162b2 were recruited to the BNT162-14 clinical trial, a Phase II, open-label, rollover trial located at multiple sites in Germany. Participants included here were vaccinated with a booster dose of BNT162b2 at least 6 months and less than 18 months since dose 2 in the parental BNT162-01 trial.

BNT162-17 is an ongoing Phase II clinical trial located at multiple sites in the US, Germany, Turkey and South Africa. Trial participants were vaccinated with BNT162b2 vaccine (30 µg, two-dose primary series) in either a clinical trial or as part of the governmental vaccination programs at least 6 months before receiving dose 3.

Participants included in this study from these trials were from subcohorts contributing to the exploratory endpoint to evaluate cross-neutralization of BNT162b2-induced antibodies to emerging SARS-CoV-2 variants following 2-dose primary series vaccination with or without booster vaccination (dose 3) in healthy adults.

The trials were carried out in accordance with the Declaration of Helsinki and Good Clinical Practice Guidelines and with approval by independent ethics committees and the competent regulatory authorities. All participants provided written informed consent. The primary objectives of these trials will be reported at a later date.

Serum specimens

Two different panels of sera were investigated. The first serum panel was obtained from 32 participants in the BNT162-01 trial drawn at a median 22 days (range 19-23 days) after receiving the second dose BNT162b2 of the 2-dose primary series (2 x 30µg). Median age was 57 years (range 20-72 years), 18 participants were ≥56 years old and none had evidence of current or prior SARS-CoV-2 at baseline. Median time from dose 1 to dose 2 was 21 days (range 19-23 days) (Table S1). The second serum panel was obtained from 30 participants in the BNT162-14 (n=11) and BNT162-17 (n=19) trial drawn at a median 28 days (range 26-30 days) after receiving the third dose BNT162b2. Median age was 44 years (range 23-72 years), 8 participants were ≥56 years old and none had evidence of current or prior SARS-CoV-2 at baseline. Median time from dose 2 to dose 3 was 219 days (range 180-342 days).

Mutation identification for Omicron

All available genome sequences assigned to the B.1.1.529 SARS-CoV-2 lineage were obtained from GISAID (*2*) on November 26th, 2021 (n=77). To retrieve the spike glycoprotein sequence from each sample, the nucleotide sequences were in-silico translated; due to ambiguity in the reading frame, each sample was translated in all three possible reading frames, and the resulting amino acid sequence was searched for the spike beginning (MFVFLVLLP) and ending (GVKLHYT). For each sample, the sequences were found in only one open reading frame. The full spike sequences were aligned using MAFFT (v7.475) (*3*) alongside with the Wuhan strain spike sequence (NCBI Reference Sequence: NC_045512.2). Two sequences were excluded due to poor alignment to the Wuhan strain. Conservation across the B.1.1.529 was used to identify the amino acid changes compared to the Wuhan strain.

VSV-SARS-CoV-2 S variant pseudovirus generation

A recombinant replication-deficient vesicular stomatitis virus (VSV) vector that encodes green fluorescent protein (GFP) and luciferase instead of the VSV-glycoprotein (VSV-G) was pseudotyped with SARS-CoV-2 spike (S) derived from either the Wuhan reference strain (NCBI Ref: 43740568), the Beta variant (mutations: L18F, D80A, D215G, R246I, Δ242–244, K417N, E484K, N501Y, D614G, A701V), the Delta variant (mutations: T19R, G142D, Δ157/158, K417N, L452R, T478K, D614G, P681R, D950N, K986P, V987P) or the Omicron variant (mutations: A67V, Δ69/70, T95I, G142D, Δ143-145, Δ211, L212I, ins214EPE, G339D, S371L, S373P, S375F, K417N, N440K, G446S, S477N, T478K, E484A, Q493R, G496S, Q498R, N501Y, Y505H, T547K, D614G, H655Y, N679K, P681H, N764K, D796Y, N856K, Q954H, N969K, L981F) according to published pseudotyping protocols (*4, 5*). In brief, HEK293T/17 monolayers (ATCC® CRL-11268™) cultured in Dulbecco’s modified Eagle’s medium (DMEM) with GlutaMAX™ (Gibco) supplemented with 10% heat inactivated fetal bovine serum (FBS [Sigma-Aldrich]) (referred to as medium) were transfected with SARS-CoV-2 S expression plasmid with Lipofectamine LTX (Life Technologies) following the manufacturer’s protocol. At 24 hours after transfection, the cells were infected at a multiplicity of infection (MOI) of three with VSV-G complemented VSVΔG vector. After incubation for 2 h at 37 °C with 7.5% CO_2_, the inoculum was removed. Cells were washed twice with phosphate buffered saline (PBS) before medium supplemented with anti-VSV-G antibody (clone 8G5F11, Kerafast Inc.) was added to neutralize residual VSV-G complemented input virus. VSV-SARS-CoV-2-S pseudotype-containing medium was harvested 20 h after inoculation, passed through a 0.2 µm filter (Nalgene) and stored at -80 °C. Prior to use in the neutralization test, the pseudovirus batches were titrated on Vero 76 cells (ATCC® CRL-1587™) cultured in medium. A defined volume of a Wuhan spike pseudovirus batch, that corresponds to an input titer of 200 transducing units (TU) per mL, was used as a reference and input volumes for the new pseudovirus batches were calculated accordingly. All pseudovirus batches used in this study were titrated simultaneously.

Pseudovirus neutralization assay

Vero 76 cells were seeded in 96-well white, flat-bottom plates (Thermo Scientific) at 40,000 cells/well in medium 4 hours prior to the assay and cultured at 37 °C with 7.5% CO_2_. Each serum was 2-fold serially diluted in medium with the first dilution of 1:5 (dilution range of 1:5 to 1:5,120). VSV-SARS-CoV-2-S particles were diluted in medium to obtain 200 TU in the assay. Serum dilutions were mixed 1:1 with pseudovirus (n=2 technical replicates per serum per pseudovirus) for 30 minutes at room temperature prior to addition to Vero 76 cell monolayers and incubation at 37 °C with 7.5% CO_2_ for 24 hours. Supernatants were removed, and the cells were lysed with luciferase reagent (Promega). Luminescence was recorded on a CLARIOstar® Plus microplate reader (BMG Labtech), and neutralization titers were calculated as the reciprocal of the highest serum dilution that still resulted in 50% reduction in luminescence. Results were reported as geometric mean titer (GMT) of duplicates. If no neutralization was observed, an arbitrary titer value of 5 (half of the limit of detection [LOD]) was reported. Tables of the neutralization titers are provided (Table S2 and Table S3).

T cell epitope conservation in the Omicron Spike variant

To estimate the rate of nonsynonymous mutation in T cell epitopes in the spike glycoprotein, we used the Immune Epitope Database (<https://www.iedb.org/>) (*6*) to obtain epitopes confirmed for T cell reactivity in experimental assays. The database was filtered using the following criteria: Organism: SARS-COV2; Antigen: Spike glycoprotein; Positive Assay; No B cell assays; No MHC assays; MHC Restriction Type: Class I; Host: Homo sapiens (human). The resulting table was filtered by removing epitopes that were “deduced from a reactive overlapping peptide pool”, as well as epitopes longer than 14 amino acids in order to restrict the dataset to confirmed minimal epitopes only. The experimental assays confirming the reactivity of these epitopes relied on multimer analysis, ELISpot or ELISpot-like assays, T cell activation assays, etc. The epitopes were reported for at least 27 different HLA-I alleles, including HLA-A, HLA-B, and HLA-C alleles. Of the 251 unique epitope sequences obtained in this approach, 244 were found in the Wuhan strain spike glycoprotein. Of these, 36 epitopes (14.8%) included a position reported to be mutated by our sequence analysis.

Statistical analysis

The statistical method of aggregation used for the analysis of antibody titers is the geometric mean and the corresponding 95% confidence interval. Using the geometric mean accounts for non-normal distribution of antibody titers that span several orders of magnitude. All statistical analyses were performed using GraphPad Prism software version 9.

**Fig. S1**

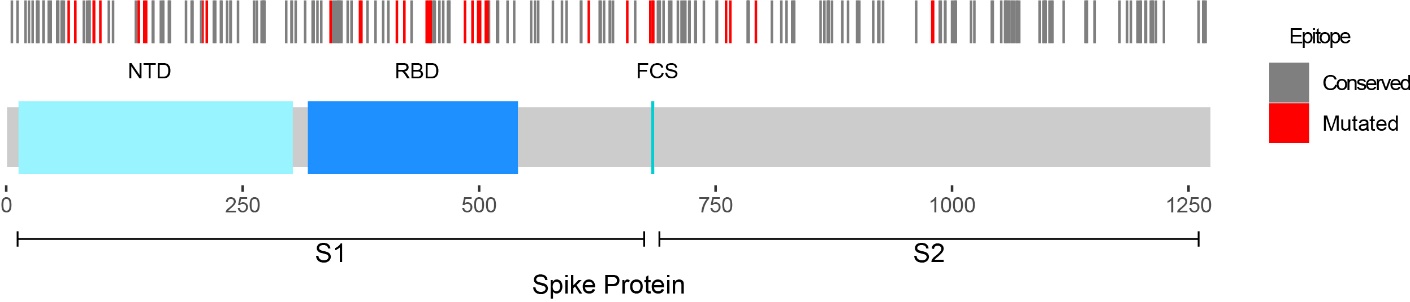
**Fig. S1. Conservation of HLA class I T cell epitopes between the Wuhan and Omicron variants.** HLA class I restricted spike protein epitopes identified based on their recognition by CD8+ T cells and reported in IEDB (n=244) are plotted by their position (top row) along the Spike protein (bottom row). Epitope indications are positioned by the amino acid position of the center of the epitope; epitopes conserved in both variants are marked in grey (n=208); epitopes spanning an Omicron mutation site are marked red (n=36). NTD=N-terminal domain; RBD=Receptor-binding domain; FCS=Furin cleavage site.

**Table S1. Cohort characteristics**

| **Characteristic** | **21d after  dose 2 cohort**  **(n=32)** | **1m after  dose 3 cohort**  **(n=30)** | **All participants  (n=51)** |
| --- | --- | --- | --- |
| Sex, n (%) |  |  |  |
| Male | 17 (53) | 15 (50) | 27 (53) |
| Female | 15 (47) | 15 (50) | 24 (47) |
| Race, n (%) |  |  |  |
| White | 32 (100) | 29 (97) | 50 (98) |
| Asian | 0 (0) | 1 (3) | 1 (2) |
| Age, median (range) | 57 (20-72) | 44 (23-72) | 46 (20-72) |
| Age group at vaccination, n (%) |  |  |  |
| 18–55 yrs | 14 (44) | 22 (73) | 33 (65) |
| 56-85 yrs | 18 (56) | 8 (27) | 18 (35) |
| Baseline SARS-CoV-2 status, n (%) |  |  |  |
| Positive | 0 (0) | 0 (0) | 0 (0) |
| Negative | 32 (100)* | 30 (100)^#^ | 51 (100)*^, #,^° |
| Unknown | 0 (0) | 0 (0) | 0 (0) |
| Interval, median (range) |  |  |  |
| Days between D1/D2 | 21 (19-23) | ‡ | n/a |
| Days until serum  draw after D2 | 22 (19-23) | n/a | n/a |
| Days between D2/D3 | n/a | 219 (180-342) | n/a |
| Days until 1m serum  draw after D3 | n/a | 28 (26-30) | n/a |

n/a = not available D = Dose, Yrs =Years, m = Month, n = Number
*, Negative SARS-CoV-2 PCR test at the time of enrollment
^#^, No evidence of prior SARS-CoV-2 infection (based on COVID-19 symptoms/signs and SARS-CoV-2 PCR test)

°, No evidence of prior SARS-CoV-2 infection in the 12 weeks prior to enrollment

‡, A subset of participants received the primary series of BNT162b2 vaccine as part of a governmental vaccination program and the interval between doses was not recorded

**Table S2. pVNT_50_ values of 32 sera collected 21 days after the second dose of 30 µg BNT162b2.**

| Clinical Study | Participant ID | pVNT_50_ | | | |
| --- | --- | --- | --- | --- | --- |
|  |  | Wuhan | Omicron | Beta | Delta |
| BNT162-01 | 2 | 160 | 5 | 10 | 40 |
|  | 6 | 320 | 5 | n/a | n/a |
|  | 3 | 80 | 5 | n/a | 80 |
|  | 10 | 160 | 10 | 40 | 80 |
|  | 11 | 320 | 10 | 320 | 320 |
|  | 12 | 160 | 5 | 20 | 80 |
|  | 9 | 160 | 5 | 20 | 80 |
|  | 7 | 320 | 20 | 80 | 160 |
|  | 13 | 160 | 5 | 40 | 40 |
|  | 14 | 160 | 10 | 20 | 80 |
|  | 15 | 160 | 5 | 10 | 80 |
|  | 5 | 160 | 5 | 10 | 40 |
|  | 16 | 160 | 5 | 160 | 80 |
|  | 4 | 80 | 5 | 10 | 40 |
|  | 17 | 160 | 5 | 20 | 80 |
|  | 18 | 640 | 10 | 40 | 160 |
|  | 19 | 160 | 10 | 80 | 160 |
|  | 20 | 160 | 5 | 20 | 80 |
|  | 8 | 20 | 5 | 5 | 5 |
|  | 21 | 160 | 5 | 10 | 80 |
|  | 22 | 40 | 5 | 5 | 10 |
|  | 23 | 80 | 5 | 5 | 40 |
|  | 24 | 160 | 10 | 20 | 160 |
|  | 1 | 80 | 5 | 20 | 40 |
|  | 25 | 160 | 5 | 20 | 80 |
|  | 26 | 320 | 10 | 40 | 160 |
|  | 27 | 80 | 5 | 20 | 40 |
|  | 28 | 80 | 5 | 10 | 40 |
|  | 29 | 320 | 20 | 40 | 160 |
|  | 30 | 320 | 10 | 40 | 160 |
|  | 31 | 640 | 20 | 40 | 160 |
|  | 32 | 320 | 20 | 80 | 160 |

n/a, not available due to lack of serum

**Table S3. pVNT_50_ values of 30 sera collected 1 month after the third dose of 30 µg BNT162b2.**

| Clinical Study | Participant ID | pVNT_50_ | | | |
| --- | --- | --- | --- | --- | --- |
|  |  | Wuhan | Omicron | Beta | Delta |
| BNT162-17 | 33 | 5120 | 1280 | 1280 | 2560 |
|  | 34 | 320 | 160 | 160 | 320 |
|  | 35 | 640 | 320 | 80 | 640 |
|  | 36 | 320 | 160 | 160 | 320 |
|  | 37 | 320 | 160 | 320 | 320 |
|  | 38 | 320 | 160 | 320 | 320 |
|  | 39 | 320 | 160 | 160 | 640 |
|  | 40 | 320 | 160 | 160 | 320 |
|  | 41 | 40 | 5 | 20 | 20 |
|  | 42 | 320 | 80 | 320 | 320 |
|  | 43 | 160 | 80 | 160 | 320 |
|  | 44 | 320 | 320 | 320 | 320 |
|  | 45 | 160 | 40 | 160 | 160 |
|  | 46 | 320 | 320 | 320 | 640 |
|  | 47 | 640 | 160 | 320 | 320 |
|  | 48 | 160 | 40 | 80 | 160 |
|  | 49 | 1280 | 640 | 640 | 1280 |
|  | 50 | 640 | 160 | 320 | 320 |
|  | 51 | 2560 | 640 | 640 | 1280 |
| BNT162-14 | 1 | 160 | 20 | 160 | 160 |
|  | 2 | 320 | 320 | 1280 | 1280 |
|  | 3 | 320 | 160 | 640 | 640 |
|  | 4 | 320 | 160 | 320 | 320 |
|  | 5 | 80 | 320 | 320 | 640 |
|  | 6 | 640 | 160 | 320 | 640 |
|  | 7 | 1280 | 1280 | 1280 | 1280 |
|  | 8 | 40 | 40 | 40 | 40 |
|  | 9 | 320 | 160 | 320 | 640 |
|  | 52 | 1280 | 640 | 1280 | 1280 |
|  | 53 | 640 | 160 | 640 | 640 |

**References**

References

1.	U. Sahin *et al.,* BNT162b2 vaccine induces neutralizing antibodies and poly-specific T cells in humans. *Nature*. **595**, 572–577 (2021), doi:10.1038/s41586-021-03653-6.

2.	S. Khare *et al.,* GISAID’s Role in Pandemic Response. *China CDC Weekly*. **3**, 1049–1051 (2021), doi:10.46234/ccdcw2021.255.

3.	K. Katoh, D. M. Standley, MAFFT multiple sequence alignment software version 7. *Molecular biology and evolution*. **30**, 772–780 (2013), doi:10.1093/molbev/mst010.

4.	M. Berger Rentsch, G. Zimmer, A vesicular stomatitis virus replicon-based bioassay for the rapid and sensitive determination of multi-species type I interferon. *PloS one*. **6**, e25858 (2011), doi:10.1371/journal.pone.0025858.

5.	A. Muik *et al.,* Neutralization of SARS-CoV-2 lineage B.1.1.7 pseudovirus by BNT162b2 vaccine-elicited human sera. *Science (New York, N.Y.)*. **371**, 1152–1153 (2021), doi:10.1126/science.abg6105.

6.	W. Fleri, N. Salimi, R. Vita, B. Peters, A. Sette, Immune Epitope Database and Analysis Resource (2016). *Encyclopedia of Immunobiology*. **2** (2016).
